# The interplay between insomnia and Alzheimer’s Disease across three main brain networks

**DOI:** 10.1101/2024.01.18.24301427

**Authors:** Jorik D. Elberse, Amin Saberi, Reihaneh Ahmadi, Monir Changizi, Hanwen Bi, Felix Hoffstaedter, Bryce A. Mander, Simon B. Eickhoff, Masoud Tahmasian, the Alzheimer’s Disease Neuroimaging Initiative

**Affiliations:** Institute of Systems Neuroscience, Medical Faculty and University Hospital Düsseldorf, Heinrich Heine University, Düsseldorf, Germany; Institute of Neuroscience and Medicine, Brain and Behavior (INM-7), Research Center Jülich, Jülich, Germany; Otto Hahn Group Cognitive Neurogenetics, Max Planck Institute for Human Cognitive and Brain Sciences, Leipzig, Germany; Faculty of Medicine, Julius-Maximilians University of Würzburg, Würzburg, Germany; Department of Neurological Diseases, Shahid Beheshti University of Medical Sciences, Tehran, Iran; Department of Psychiatry and Human Behavior, University of California, Irvine, CA 92603, USA; Department of Nuclear Medicine, University Hospital and Medical Faculty, University of Cologne, Cologne, Germany

**Keywords:** Insomnia, Alzheimer’s disease, Mild Cognitive Impairment, Default Mode Network, Salience Network, Central Executive Network

## Abstract

**Background:** Insomnia is prevalent along the trajectory of Alzheimer’s disease (AD), but the neurobiological underpinning of their interaction is poorly understood. Here, we assessed structural and functional brain measures within and between the default mode network (DMN), salience network (SN), and central executive network (CEN).

**Methods:** We selected 320 subjects from the ADNI database and divided by their diagnosis: cognitively normal (CN), Mild Cognitive Impairment (MCI), and AD, with and without self-reported insomnia symptoms. We measured the gray matter volume (GMV), structural covariance (SC), degrees centrality (DC), and functional connectivity (FC). We tested the effect and interaction of insomnia symptoms and diagnosis on each index across groups. Subsequently, we performed a within-group linear regression for each network. Finally, we correlated brain abnormalities with cognitive scores.

**Results:** Insomnia symptoms were associated with FC alterations across all groups. The AD group also demonstrated an interaction between insomnia and diagnosis. In the CN and MCI groups, insomnia symptoms were characterized by patterns of within-network hyperconnectivity, while in the AD group, within- and between-network hypoconnectivity was observed. FC alterations within and between DMN and CEN hubs were associated with reduced MMSE scores in each of the groups. SC and GMV alterations were non-significant in the presence of insomnia symptoms, and DC indices only showed network-level alterations in the CEN for AD individuals.

**Conclusions:** Insomnia symptoms affect the intrinsic functional organization of the three networks along the AD trajectory. Patients with AD present with a unique pattern of insomnia-related functional alterations, highlighting the profound interaction between both conditions.

## INTRODUCTION

Sleep disturbances are a common catalyst and consequence of Alzheimer’s disease (AD) pathology. This is of critical importance, as 40 to 70% of older adults at risk for AD reported chronic sleep problems,^1^ and sleep impairments affect at least 24% of patients with mild to moderate AD and 39% of patients with severe AD.^2,3^ Animal and human studies demonstrated that in the prodromal and preclinical stages of AD, sleep disturbances induce higher beta-amyloid burden in the brain,^4–7^ enhanced tau accumulation in the interstitial fluid (ISF) and cerebrospinal fluid (CSF),^4^ increased risk of clinical diagnosis of AD,^8^ and accelerated cognitive decline.^2,8–14^ In the clinical stage, sleep disturbances may be exacerbated by (tau-driven) deterioration of brain regions/networks involved in sleep and circadian rhythm,^15^ the emergence of abnormal sleep-wake behaviors, and certain types of medication.^13^ Hence, there is bidirectional reinforcement between sleep disturbances and AD pathophysiology^16^ that remains to be further disentangled using large-scale cohorts.

Of all possible causes for sleep disturbances, including sleep-disordered breathing, restless leg syndrome, and narcolepsy, insomnia symptoms are among the most prevalent, affecting nearly 30-35% of adults.^17,18^ This is due in part to their high comorbidity with neurological and affective disorders.^19,20^ Insomnia symptoms are characterized by perceived difficulties in falling asleep, maintaining sleep, and early waking times, not attributable to sleep-disrupting external conditions and accompanied by subjective daily dysfunctions.^21^ Chronic insomnia disorder (ID), in turn, is characterized by the occurrence of these symptoms at least thrice a week and for a period of at least three months according to the International Classification of Sleep Disorders-third Edition (ICSD-3).^22^ In spite of the high prevalence of both insomnia symptoms and chronic insomnia disorder in middle-aged and older subjects, the neurobiological correlates of insomnia remain relatively poorly understood. Multimodal neuroimaging studies have suggested that insomnia may primarily be a functional brain disorder, that is to say, caused by aberrant connectivity of certain brain regions/networks.^18^

A recent study based on 29,423 subjects found that insomnia symptoms were associated with increased functional connectivity (FC) within the default mode network (DMN), salience network (SN), and central executive network (CEN, also known as the frontoparietal network or FPN), as well as decreased FC between the DMN and SN, and between the DMN and CEN.^23^ These three intrinsic brain networks, are primarily involved in self-referential processing, mind-wandering, rumination (DMN), detecting emotional components of external and interoceptive stimuli, switching between mind-wandering and directed thought (SN), and working memory, decision-making, and goal-directed behavior (CEN) comprise the triple network model,^24^ which has been studied in the context of various neuropsychiatric disorders, including AD.^25–28^ Indeed, the DMN is often an early target of AD-related pathological changes such as amyloid-β and tau accumulation, altered glucose metabolism, abnormal FC, and brain atrophy.^29–32^ These phenomena, particularly tau accumulation, spread along other connected brain regions/networks.^28,33,34^ It has been suggested that AD pathology within these networks may be a driving factor behind the development of insomnia symptoms in many AD individuals due to tau-driven degeneration of sleep-regulating neurons.^15,35^ Taken together, the interrelationship between insomnia and AD may be anchored in the DMN, SN, and CEN, which warrants further exploration of potential structural and functional alterations within and between these three networks. Although it is clear that insomnia and AD independently affect the internal connectivity and morphology of these networks, the present study aims to investigate whether insomnia symptoms modulate the extant functional and structural impairments along the trajectory of AD, and whether this modulatory effect could be correlated with cognitive decline.

In this pre-registered study (https://osf.io/2bju9), we investigated the role of insomnia symptoms on the triple-network system along the trajectory of AD (cognitively normal (CN), patients with MCI and AD) using the Alzheimer’s Disease Neuroimaging Initiative (ADNI) database. In particular, we assessed how the presence of insomnia symptoms correlated with four MRI indices relating to both the connectivity and morphology of the three brain networks FC, degrees centrality (DC), structural covariance (SC), and gray matter volume (GMV). These indices reflect the degree of functional co-activation between nodes, the degree of involvement in co-activation pairs of individual nodes, the degree of structural co-association of nodes reflected by shared alterations in gray matter, and the changes in gray matter of individual nodes, respectively, providing a comprehensive insight into the activity and integrity of the three networks.

## MATERIALS AND METHODS

### Participants

The Alzheimer’s Disease Neuroimaging Initiative (ADNI) is a collaboration of 63 research sites across the US and Canada to collect, validate, and utilize AD data, including MRI and PET images, genetic assays, CSF and blood biomarkers, and cognitive assessments.^36^ The ADNI database (adni.loni.usc.edu) contains more than 2000 individuals subdivided into elderly control, early MCI, late MCI, and clinical AD groups. The present study uses data from the second and third phases of the initiative (ADNI-2 and ADNI-3), with collection dates ranging from 2011 to 2021. For this study, 1646 individuals were selected based on the availability of structural and functional MRI scans and a neuropsychiatric inventory (NPI) assessment of insomnia symptoms. We excluded subjects if there was a change in diagnosis of patients within six months of the NPI or if the imaging data collection had occurred longer than six months from the NPI. We also excluded individuals who had not performed a mini-mental state exam (MMSE) or geriatric depression scale (GDS) test, who obtained a score of 9 or higher on the GDS (indicating comorbid depressive symptoms), or who were diagnosed with sleep disorders other than insomnia, including restless leg syndrome, obstructive sleep apnea, or narcolepsy. After selecting eligible subjects, they were grouped into three diagnoses (CN, MCI, and AD). Within these groups, individuals with insomnia symptoms (+insomnia) and individuals without (-insomnia) were carefully matched using propensity scores taking into account the individuals’ age, sex, handedness, education, MMSE and GDS scores, yielding a matched raw sample of 354 (**Figure 1, Table 1**).

**Figure 1.**
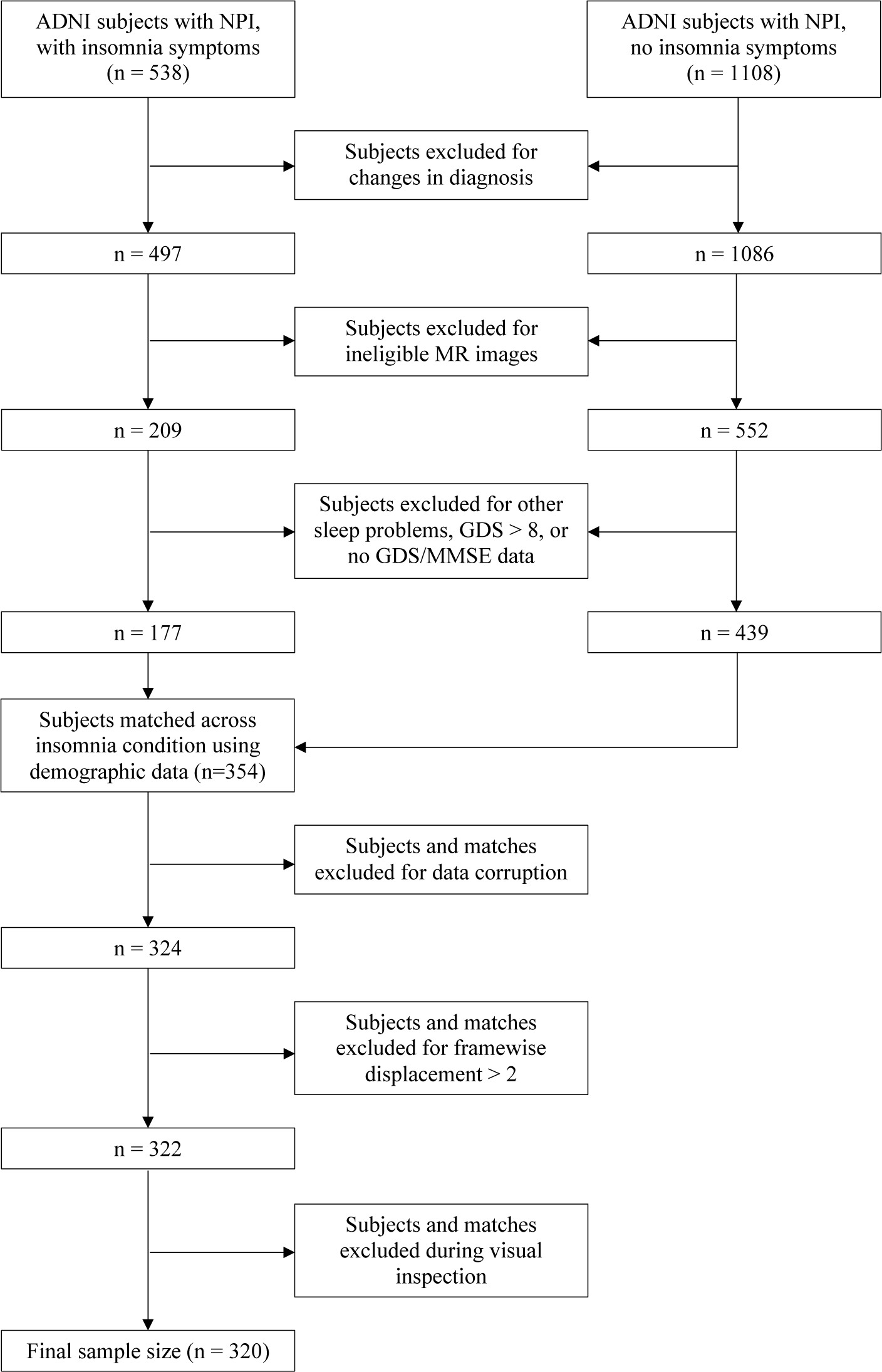
– Flowchart of Participant Selectionss. Inclusion and exclusion procedures before and after subject matching are depicted. *GDS = Geriatric Depression Scale; MMSE = Mini Mental State Exam; NPI = Neuropsychiatric Inventory*.

**Table 1.** - Participant Demographics. Demographic variables are presented as counts with percentages (Total N, Sex) or as means with standard deviations (Age, Education, MMSE, and GDS). *GDS = Geriatric Depression Scale; MMSE = Mini Mental State Exam. *p < 0.05, **p < 0.01, ***p < 0.001, ****p < 0.0001. ß = estimate*

|  | Main Dataset (N = 320) |  |  |
| --- | --- | --- | --- |
|  | -Insomnia | +Insomnia | p-value |
| **Cognitively Normal (CN)** |  |  |  |
| Total N | 74 (50%) | 74 (50%) |  |
| Age | 73.536 (7.8) | 74.412 (7.3) | 0.481 |
| Sex (Male/Female) | 21 (28%) / 53 (72%) | 20 (27%) / 54 (73%) | 1.000 |
| Years of Education | 16.270 (2.5) | 16.432 (2.3) | 0.677 |
| MMSE | 28.905 (1.2) | 29.081 (1.2) | 0.376 |
| GDS | 1.149 (1.4) | 1.297 (1.4) | 0.520 |
| **Mild Cognitive Impairment (MCI)** |  |  |  |
| Total N | 66 (50%) | 66 (50%) |  |
| Age | 72.558 (7.5) | 73.621 (7.8) | 0.424 |
| Sex (Male/Female) | 43 (65%) / 23 (35%) | 31 (47%) / 35 (53%) | 0.054 |
| Years of Education | 15.712 (2.8) | 15.530 (2.6) | 0.704 |
| MMSE | 28.136 (1.7) | 27.985 (1.8) | 0.619 |
| GDS | 2.030 (1.8) | 2.288 (2.0) | 0.438 |
| **Alzheimer's Disease (AD)** |  |  |  |
| Total N (%) | 20 (50) | 20 (50) |  |
| Age | 77.240 (8.1) | 77.685 (6.7) | 0.851 |
| Sex (Male/Female) | 10 (50%) / 10 (50%) | 10 (50%) / 10 (50%) | 1.000 |
| Years of Education | 15.400 (2.4) | 15.700 (2.6) | 0.706 |
| MMSE | 21.650 (3.2) | 21.050 (5.5) | 0.675 |
| GDS | 1.600 (1.7) | 1.950 (1.7) | 0.523 |

### Insomnia Symptoms

The subsection K (“Sleep”) of the NPI^37^ was used to identify whether subjects had experienced insomnia symptoms around the time their images were taken. Specifically, if “yes” had been answered to either questions K1 (“Does the patient have difficulty falling asleep”), K2 (“Does the patient get up during the night (do not count if the patient gets up once or twice per night to go to the bathroom and falls back asleep immediately)?”), or K6 (“Does the patient awaken too early in the morning (earlier than was his/her habit)?”), individuals were considered as having insomnia symptoms (+insomnia). This method has been successfuly used in numerous AD studies to assess the presence of insomnia.^38^

### Imaging Data

MRI acquisition protocols have been described in detail elsewhere.^36^ For each subject, the 3T T1-weighted (T1w) structural MRI and resting state functional MRI (rs-fMRI) were obtained from the online ADNI database. The native DICOM files were converted into the BIDS format^39^ using a custom script based on dcm2niix. BIDS verification was done by the validator within fMRIPrep.^40^ After verification, the raw data was stored as a DataLad^41^ dataset for easy management and version control. The T1w and rs-fMRI images were preprocessed using the standard fMRIPrep pipeline, with additional motion correction performed by ICA-AROMA for functional data.^42^ Using Nilearn as part of a custom script, we subsequently removed the first four volumes of the rs-fMRI image, detrended and filtered the image (0.01 – 0.1 Hz bandpass), regressed out the 24 motion parameters obtained from the fMRIPrep confounds, and smoothed the image at 5 mm FWHM. As a final preprocessing step, all anatomical and functional scans were manually inspected. Subjects and their matches were excluded if the anatomical or functional images were revealed to be corrupted (e.g. containing missing data) during the fMRIPrep preprocessing, if the framewise displacement of the functional images exceeded 2 degrees, or if the anatomical or functional image did not pass manual inspection (**Figure 1**). All selection and exclusion processes yielded a final matched sample of 320.

### Functional Connectivity

The whole-brain fMRI time series of each subject was extracted using Nilearn. Nodes within the triple network model were identified using the 100-parcel, 7-network Schaefer atlas.^43^ The bilateral nodes of the SN (n = 12), DMN (n = 24), and CEN (n = 13) were isolated and their time series stored and sorted by the network. These time series were used to create a 49 x 49 correlation matrix assessing the FC between all within- and between-network nodes within the triple-network system.

### Degrees Centrality

The DC was calculated by binarizing the FC matrix using three different absolute thresholds (0.2, 0.4, and 0.6) and counting the number of connections to each node to obtain the nodal DC for each threshold.

### Gray Matter Volume

Region-based morphometry (RBM) for gray matter volume was performed on the preprocessed T1w images using the standalone version of the CAT12 toolbox and the 100-parcel, 7-network Schaefer atlas. CAT offers the capability to perform regional analyses using RBM, in addition to voxel-wise analyses like voxel-based morphometry (VBM).^44^ Our RBM pipeline comprised three steps: tissue segmentation of the T1w scans, registration to MNI space, and extraction of RBM from the 100 Schaefer parcellations within each subject’s native brain space by calculating the mean over gray matter voxels for bilateral nodes in the three networks.

### Structural Covariance

SC matrices were obtained for each diagnostic group, and different insomnia conditions within each group by correlating mean GMV between individual three network nodes. These matrices assessed the degree to which morphological changes of one node, i.e. changes in GMV, were coupled to morphological changes in another node.^45^ These correlations were corrected for age and sex.

### Statistical Analysis

Descriptive statistical data are presented as mean values with standard deviations. To test whether insomnia symptoms have a significant effect on the three network indices, we fit a linear regression model for each index taking the index as the dependent variable, the (interaction between) insomnia and diagnosis as the independent variable, and age, sex, framewise displacement and gray matter volume as covariates-of-no-interest. After fitting these models, we performed a series of cross-sectional tests.

To investigate the relationship between insomnia symptoms and FC alterations at several stages along the trajectory of AD, we fit a series of linear regression models correlating insomnia symptoms to changes in aggregate FC categorized by networks or 25 regions-of-interest (ROIs). First, we fit one model on all FC values across all diagnostic groups to assess the interaction between insomnia and diagnosis. We fit 18 models with insomnia symptoms as the independent variable and the aggregate FC values of all edges belonging to a specific within- or between-network category (i.e. all edges between the DMN and SN) as the dependent variable. We additionally fit 942 models with insomnia symptoms as the independent variable, and the aggregate FC values of all edges belonging to a specific within- or between-ROI category (i.e. all edges between the left parietal DMN and right medial SN) as the dependent variable. These models were corrected for age, sex, framewise displacement, and total GMV. The estimates (ß) of these models were entered into heat maps representing all network- and ROI-level edges within the triple-network system. To investigate the relationship between insomnia symptoms and DC/GMV, we fit an additional 196 models in order to cover all 49 nodes and each DC threshold (0.2, 0.4, 0.6), corrected for age and sex. The model-specific assumption of linearity was met on account of the binary classification of the independent variable (-insomnia vs. +insomnia). The assumption of homoscedasticity was met in 97.5% of tests, and the assumption of normality in 57.5% of tests. Both percentages were considered acceptable for this study, due to the negligible prevalence of heteroscedastic residuals and the minimum number of observations (n = 320) being large enough to fall under the Central Limit Theorem.

To investigate the effect of insomnia symptoms on SC matrices along the trajectory of AD, we performed permutation tests across different diagnostic groups and insomnia conditions. This was done by shuffling the -insomnia and +insomnia labels one thousand times and calculating the mean difference to create an approximate statistical distribution, allowing for an estimation of the probability of observed (actual) mean differences for each comparison pair.

The correlations between FC alterations and MMSE score were obtained by performing an age- and sex-corrected linear regression using the ROI-level indices as predictor variables, chosen because they exhibited the most significant changes in prior analyses. For all statistical analyses, we took p<0.05 as indicating significance, after performing the false discovery rate (FDR) correction for multiple comparisons.^46^

## RESULTS

### Participant Demographics

The main sample (n = 320) consisted of 135 male and 185 female participants with a mean age of 74.0 years (SD = 7.64). Of these participants, 148 were CN, 132 were diagnosed with MCI, and 40 with AD. Half the participants (n = 160) reported insomnia symptoms. Sociodemographic and clinical differences across the -insomnia and +insomnia conditions are displayed in **Table 1**.

### Aberrant Functional Connectivity Associated with Insomnia Symptoms

Insomnia symptoms were associated with significant FC alterations across all nodes and diagnoses (p < 0.01). Moreover, there was a significant interaction between insomnia and diagnosis for AD individuals (p < 0.001), but not CN and MCI individuals. Within diagnostic groups, insomnia symptoms were associated with varying patterns of FC abnormalities, both at the level of networks and ROIs. In CN and MCI, +insomnia was characterized by patterns of increased FC within networks, while in AD, +insomnia was characterized by patterns of decreased FC within and between networks (**Figure 2**). Within-DMN FC alterations were significantly associated with insomnia symptoms across all groups. In CN and MCI, within-DMN connectivity was increased, driven by enhanced co-activation of the bilateral precuneus and the left prefrontal cortex (PFC). In AD, within-DMN connectivity was decreased, largely due to attenuated co-activation of the bilateral temporal DMN and left parietal DMN. Interestingly, all within-DMN edges affected in CN/MCI +insomnia was non-significant in AD +insomnia, and vice versa. This suggests that within-DMN ROIs are differentially affected in their (co-)activity by insomnia symptoms along the trajectory of AD.

**Figure 2.**
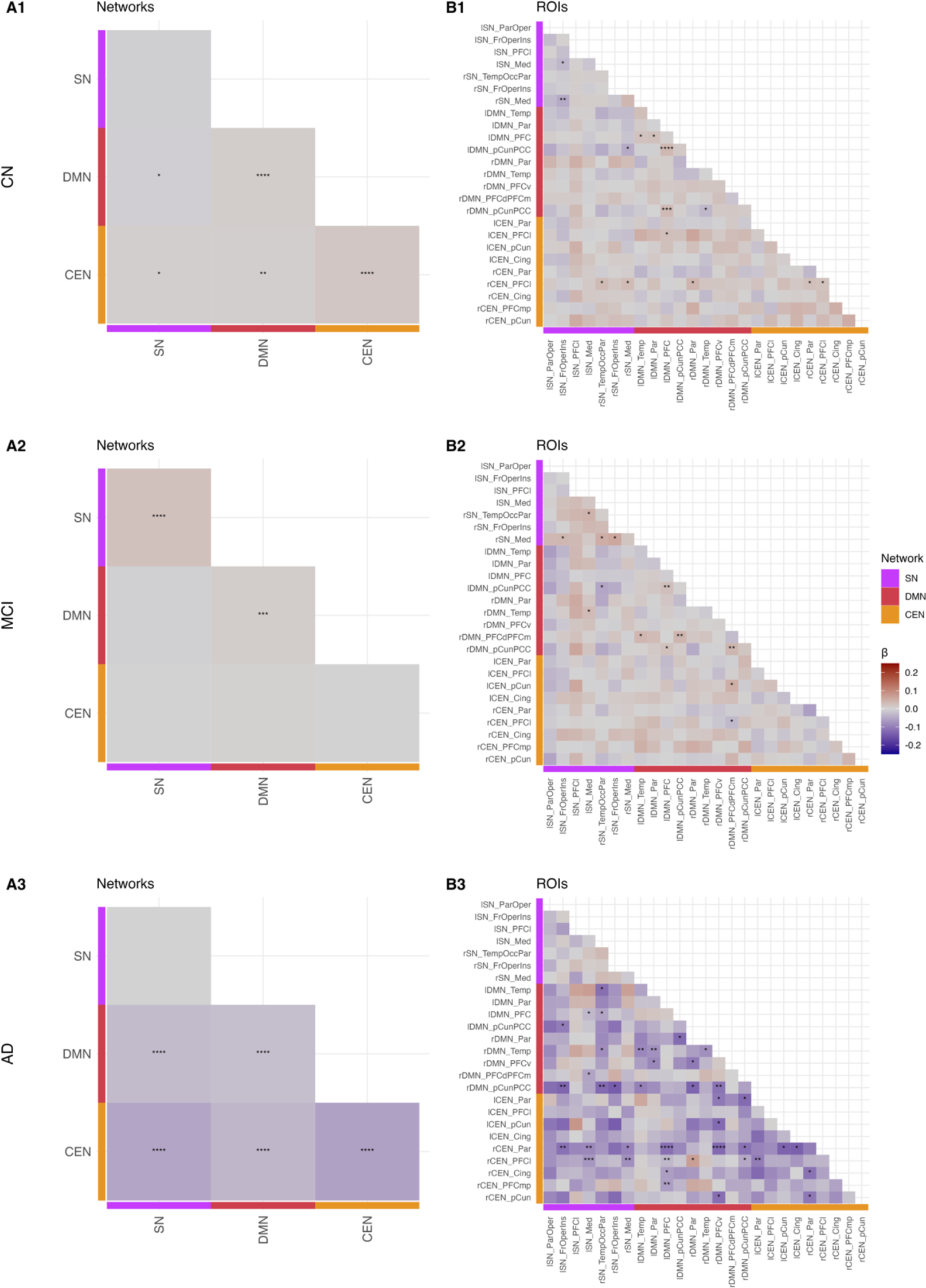
– Three Network Functional Connectivity Changes in the Presence of Insomnia Symptoms. (**A)** Network- and (**B**) ROI-level alterations in three network connectivity associated with insomnia symptoms in the CN, MCI, and AD groups. *CEN = Central Executive Network; Cing = cingulate; DMN = Default Mode Network; Fr = frontal; Ins = insula; Med = medial; Occ = occipital; Oper = operculum; Par = parietal; PCC = posterior cingulate cortex; pCun = precuneus; PFC = prefrontal cortex; PFCd = dorsal prefrontal cortex; LPFC = lateral prefrontal cortex; PFCm = medial prefrontal cortex; PFCmp = medial posterior prefrontal cortex; PFCv = ventral prefrontal cortex; SN = Salience Network; Temp = temporal. *p < 0.05, **p < 0.01, ***p < 0.001, ****p < 0.0001. ß = estimate*.

FC alterations within-CEN, CEN-SN, and CEN-DMN were prevalent in the CN and AD groups, but not in MCI. As within the DMN, these CEN edges were hyperconnective in CN and hypoconnective in AD. In CN, increased FC was driven by enhanced co-activation of the right lateral PFC (LPFC) with various hubs of the three networks. In AD, decreased FC was associated with attenuated co-activation of the bilateral LPFC, right parietal CEN, and various hubs of the three networks. Overall, whereas the connectivity profile of the CN group showed increased modulation by the CEN, the findings in the AD group suggested a widespread dysconnectivity of the CEN from the other two networks. Unlike within the DMN, these FC alterations largely centered around the same ROIs, namely the LPFC.

Within-SN and SN-DMN alterations were a significant marker of insomnia in the MCI group and were present to a lesser extent in the CN and AD groups. Increased within-SN connectivity in MCI was driven by enhanced correlation of the frontal operculum, insula, and medial SN. These same ROIs exhibited various amounts of attenuation in their correlation with DMN hubs, including the precuneus and PFC, across all diagnostic groups. This reduction in SN-DMN FC was the only consistent correlate of insomnia symptoms that was evident across the entire trajectory of AD.

### Aberrant Network-Level Degrees Centrality Associated with Insomnia Symptoms

Insomnia symptoms were associated with significant DC alterations across all diagnostic groups for the 0.4 absolute threshold (p < 0.01), but not the 0.2 and 0.6 thresholds, indicating a dependency on thresholding. Within-group analyses revealed non-significant DC changes of individual nodes or ROIs associated with insomnia symptoms (**Figure S5A**) for all thresholds. Aggregate DC alterations at the network-level were non-significant for the DMN and SN, but CEN DC values were reduced in AD +insomnia (p < 0.05) at all thresholds. This is in line with the significant CEN dysconnectivity identified in AD in the within-group FC analyses.

### No Structural Covariance and Gray Matter Volume Alterations Associated with Insomnia Symptoms

Permutation tests revealed that the three network SC did not significantly change between the insomnia conditions across diagnostic groups (**Figure S4**). Furthermore, the +insomnia condition was not associated with significant alterations in the GMV of main nodes of the three networks (**Figure S5B**).

### Functional Connectivity Abnormalities Associated with Cognitive Decline

To investigate whether the observed FC abnormalities in CN/MCI/AD individuals with insomnia symptoms were associated with cognitive decline, we tested the effect of aberrant ROI-level FC on the MMSE scores of individuals. Within the CN and MCI groups, within-DMN hyperconnectivity, especially increased FC between the PFC and precuneus, appeared to be the most significant correlate of reduced cognitive function (**Figure S6, Figure 3**). In AD, several hypoconnective within-DMN, DMN-SN, and SN-CEN edges were associated with reduced cognitive function (**Figure 4**). A single SN-CEN edge, between the medial SN and LPFC, was also associated with higher cognitive function. These findings indicate that major FC alterations in each of the groups can be associated with cognitive decline and, crucially, that both hyperconnectivity (as seen in e.g. the DMN of CN individuals) and hypoconnective (as seen in e.g. the DMN of AD individuals) alterations can be detrimental.

**Figure 3.**
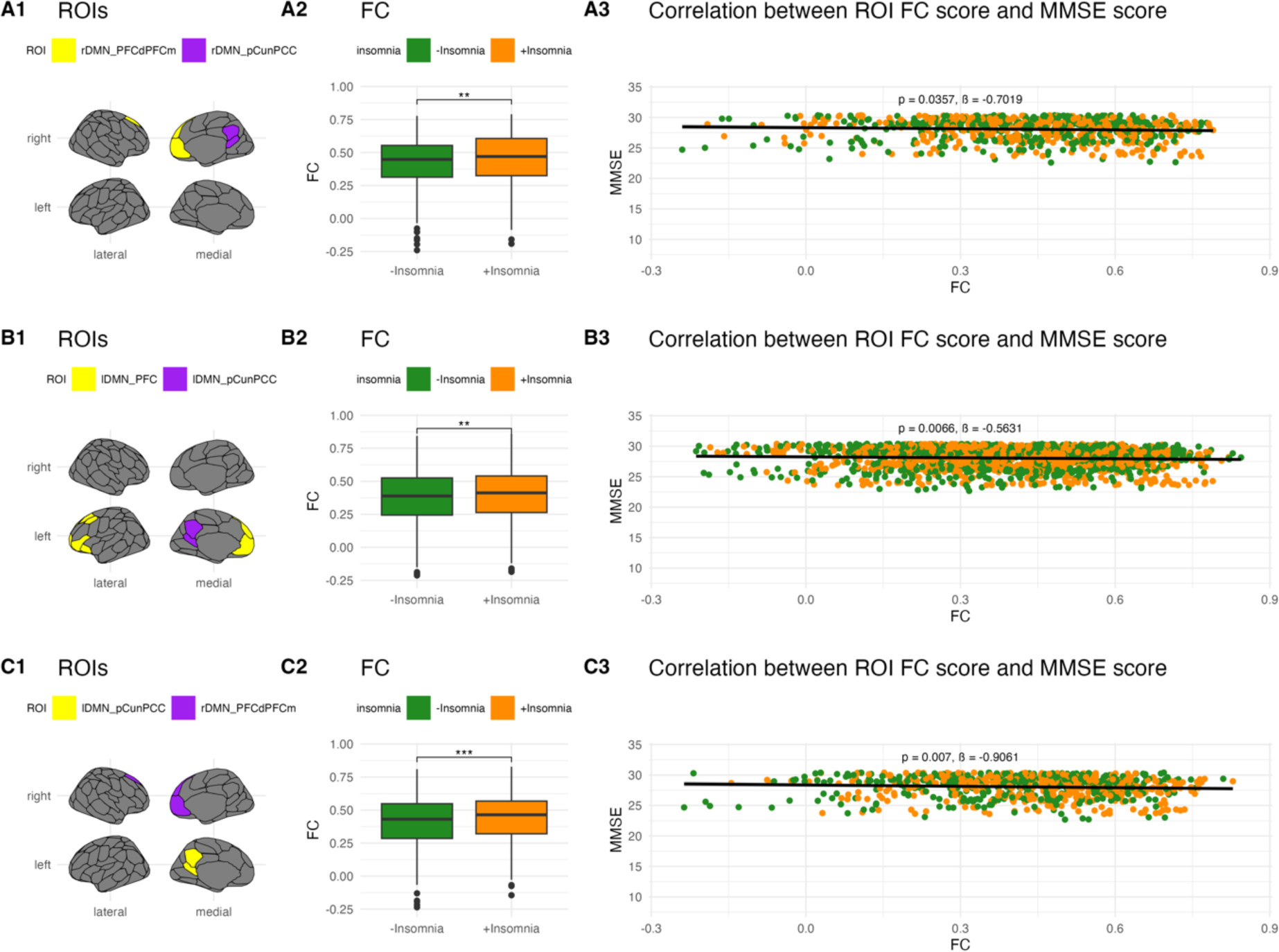
- Abnormal Inter-ROI Connectivity and MMSE Score in MCI. ROI-level edges in MCI +insomnia that predict significant changes in MMSE score. (**1**) ROI-level edges affected by the +insomnia condition. (**2**) Change in FC for selected edges across the +insomnia condition. (**3**) Overall correlation of ROI-level edge FC scores and MMSE scores. Hyperconnective intra-DMN edges are overwhelmingly associated with decreased cognitive functioning in MCI +insomnia. *DMN = Default Mode Network; FC = functional connectivity; PCC = posterior cingulate cortex; pCun = precuneus; PFC = prefrontal cortex; PFCd = dorsal prefrontal cortex; LPFC = lateral prefrontal cortex. *p < 0.05, **p < 0.01, ***p < 0.001, ****p < 0.0001. ß = estimate*.

**Figure 4.**
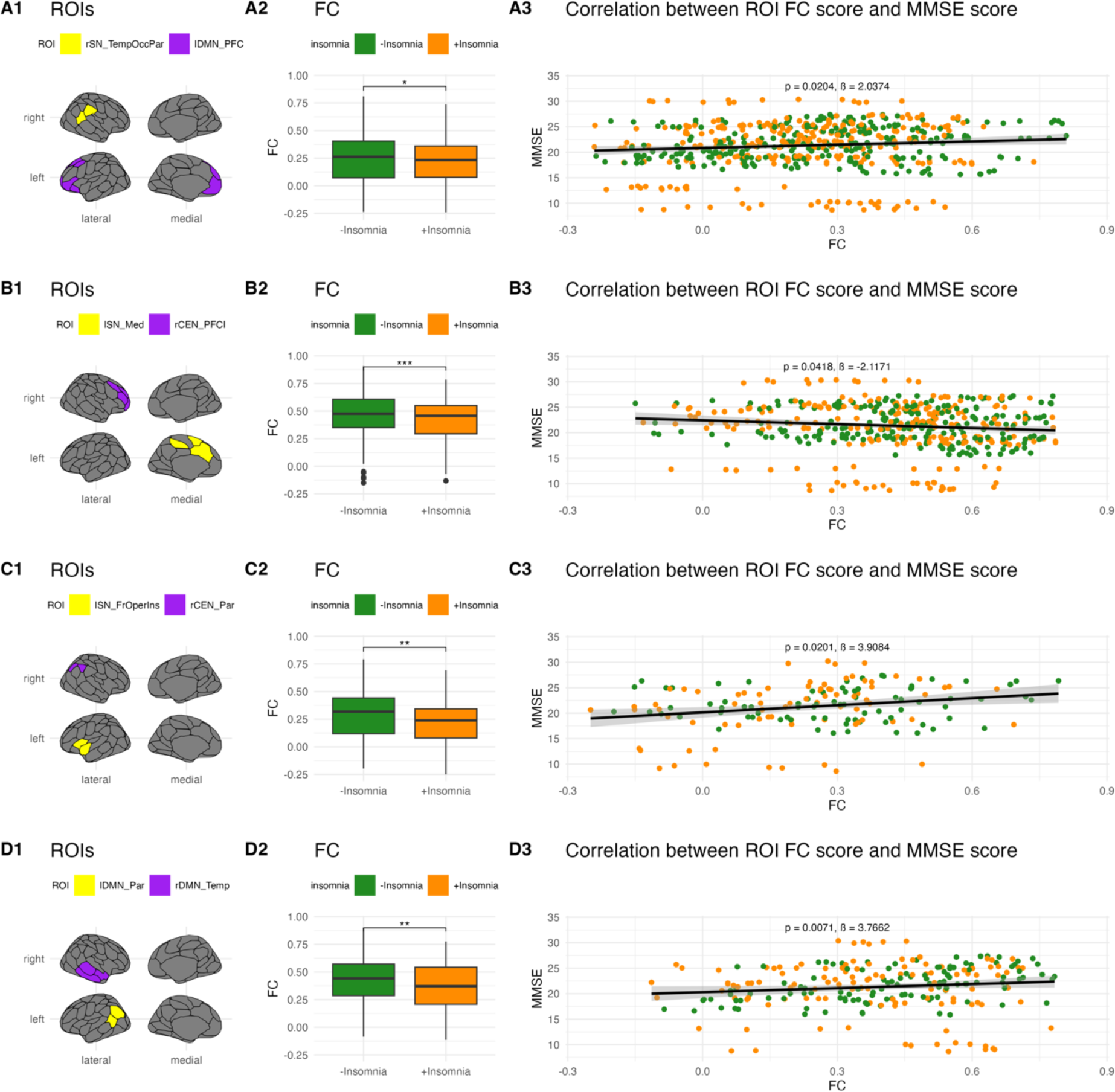
- Abnormal Inter-ROI Connectivity and MMSE Score in AD. ROI-level edges in AD +insomnia that predict significant changes in MMSE score. (**1**) ROI-level edges affected by the +insomnia condition. (**2**) Change in FC for selected edges across the +insomnia condition. (**3**) Overall correlation of ROI-level edge FC scores and MMSE scores. Certain hypoconnective intra-DMN, inter-SN-DMN, and inter-SN-CEN edges predicted decreased cognitive functioning in AD +insomnia. One hypoconnective inter-SN-CEN edge predicted increased cognitive functioning in AD +insomnia. *CEN = Central Executive Network; DMN = Default Mode Network; FC = functional connectivity; Fr = frontal; Ins = insula; Med = medial; Occ = occipital; Oper = operculum; Par = parietal; PFC = prefrontal cortex; LPFC = lateral prefrontal cortex; SN = Salience Network; Temp = temporal. *p < 0.05, **p < 0.01, ***p < 0.001, ****p < 0.0001. ß = estimate*.

## DISCUSSION

The present study investigated the effect of insomnia symptoms on four functional and structural indices of the triple-network system along the trajectory of AD. We found that FC was the only index significantly affected by insomnia symptoms across all diagnostic groups and all levels of analyses. We also found an interaction between the effect of insomnia symptoms and diagnosis in the AD group. Cross-sectional, within-group analyses of major components of the three networks revealed patterns of increased FC associated with insomnia symptoms in the CN and MCI groups, and patterns of decreased FC in the AD group. The CN and MCI groups exhibited within-DMN, within-SN (MCI only), and CEN hyperconnectivity, whereas the AD group exhibited within-DMN and CEN hypoconnectivity. In contrast, all groups exhibited some degree of SN-DMN hypoconnectivity. These results were not reflected by similar DC alterations. Only the aggregate CEN DC of the AD group was significantly reduced at all thresholds. The SC and GMV were not significantly affected by insomnia symptoms across the trajectory of AD. A summary of the main findings of this study is depicted in **Figure 5**.

**Figure 5.**
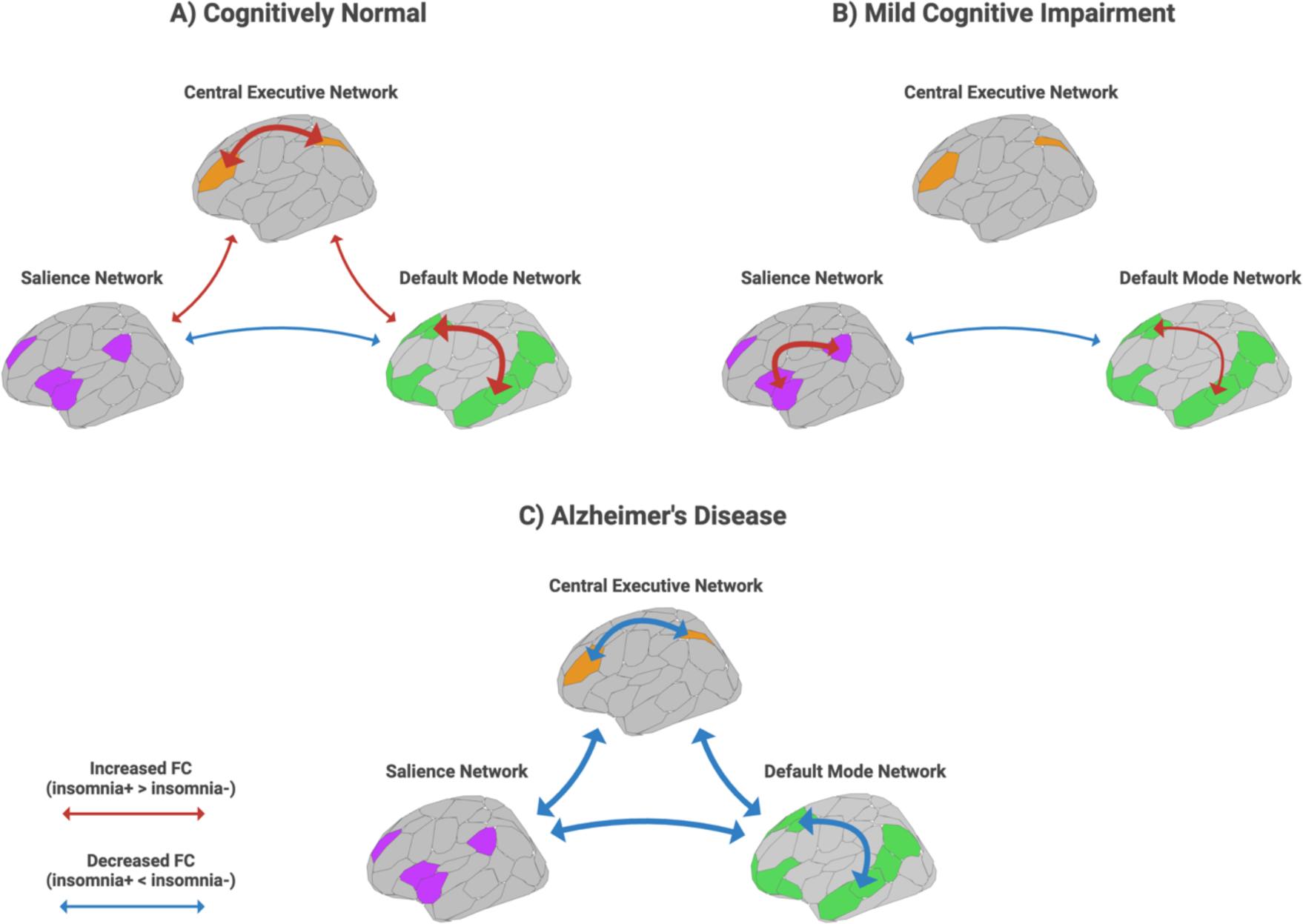
– Summary of Main Findings. Summary of network alterations associated with insomnia symptoms along the trajectory of AD. Within- and between-network FC alterations were the only significant marker of insomnia across the (**A**) Cognitively Normal (**B**) Mild Cognitive Impariement, and (**C**) Alzheimer’s disease groups. Arrow width represents relative strength of FC alterations. *FC = functional connectivity*.

### Interaction Between Alzheimer’s Disease and Insomnia Symptoms

AD pathophysiology is typically associated with both structural and functional alterations^47,48^ spreading along networks like the DMN, SN, and CEN.^27,28,49^ In contrast, functional brain network alterations, but not structural alterations, are most often found to underlie insomnia symptoms.^23,50,51^ In line with this, we found that insomnia symptoms only affected FC and, to a lesser extent, DC of the three networks. Interestingly, the patterns of hyperconnectivity identified in the CN and MCI group mimicked the connectivity alterations typically found in insomnia,^23^ whereas those in AD did not. To wit, within-DMN FC alterations are typically seen as a correlate of hyperarousal in insomnia, and precuneus-PFC hyperconnectivity was observed in individuals with insomnia symptoms in the CN and MCI groups. Insomnia symptoms in AD were instead associated with intratemporal and temporal-parietal hypoconnectivity. However, both patterns of FC alterations were associated with decreased cognitive function. Similarly, LPFC connectivity, which reflects the degree of DMN regulation by the CEN, was increased in CN but decreased in AD in the presence of insomnia symptoms. All of the above suggests that insomnia symptoms and AD have a profound, interdependent effect on the functional organization of the three networks.

### DMN Connectivity Alterations and Associated Symptoms

Current knowledge of the triple-network system suggests that even minor FC alterations of its interdependent components may be associated with major pathological manifestations (B. Menon, 2019). In the context of both insomnia and AD, three network dysfunctions are broadly correlated with deficits in attention and cognitive control, abnormal arousal states, and maladaptive thought and behavior.^23,26,52^ Baseline DMN activity guides self-referential mental processes at rest, whereas hyperactivity may accompany more maladaptive forms of self-referential thought, such as rumination. Indeed, DMN hyperactivity is an important finding in insomnia, as it is frequently associated with hyperarousal states at the onset of sleep.^53^ We identified a significant within-DMN hyperconnectivity in both CN and MCI, characterized by an increased FC between the precuneus/PCC and the DMN-associated PFC, two main hubs of the DMN.^54^ DMN hyperactivity may sometimes be a result of inadequate downregulation by the SN, i.e. the inadequate suppression of mind wandering and rumination when required.^55^ Indeed, we found signs of reduced DMN-SN connectivity in CN and MCI as well, intersecting at the precuneus/PCC. Curiously, there was also evidence of reduced DMN-SN connectivity in AD, but without DMN hyperconnectivity. A possible explanation could be the degeneration of precuneus-centric subnetworks, as the structure is prone to high levels of Aß accumulation.^29^ This could lead to decreased baseline connectivity, meaning that, even in the case of inadequate regulation by the SN, the subnetwork would not become hyperconnective. Indeed, we do see that many hyperconnective edges in CN/MCI are non-significant in AD. However, this does not fully explain the prominent within-DMN hypoconnectivity we see in AD +insomnia. One explanation could be exacerbated functional disintegration of the DMN, as the DMN is an early target of AD pathophysiology^56^ and insomnia symptoms may increase the severity of neurodegeneration.^57^ However, DMN hypoactivity has also been reported in insomnia as a correlate of emotional dysregulation and reduced daytime attentiveness as a consequence of poor quality of sleep.^50,58,59^ We speculate that extant comorbidities of progressed AD may specifically exacerbate the affective symptoms of insomnia, leading to the significant link between temporal-parietal DMN dysconnectivity and insomnia symptoms in this group.

### SN Connectivity Alterations and Associated Symptoms

The SN is predominantly involved in salience detection and arousal, as well as regulating the activity of the DMN and CEN in concert.^55^ In insomnia, SN hyperconnectivity may be correlated with increased arousal as a result of sustained processing of environmental and proprioceptive stimuli.^50,60^ Conversely, SN hypoconnectivity is not a common phenomenon in insomnia but has been found to correlate with negative affect and apathy in late-life depression.^61^ We found that, in the MCI group, insomnia symptoms were associated with significantly increased connectivity of the medial SN with the bilateral frontal SN, the operculum, and the insula. Insular connectivity aberrations are consistently associated with various neuropsychiatric conditions, including insomnia.^62^ Although within-SN hyperconnectivity has been implicated in many sleep studies as a correlate of emotional hyperarousal, many studies may have inadequately controlled for depression and anxiety levels.^50,63,64^ Crucially, we did not find that within-SN hyperconnectivity predicted a decreased MMSE in MCI +insomnia, suggesting these connectivity alterations are not as impactful on daytime function as the ones identified in the DMN.

### CEN Connectivity Alterations and Associated Symptoms

The CEN is involved in working memory and goal-oriented action and is critically affected in anxiety disorders.^55^ CEN hyperconnectivity has been suggested to reflect a compensatory mechanism for sleep disturbance, as it inhibits emotional dysregulation and facilitates simpler cognitive processes that are inhibited by poor sleep.^65^ CEN hypoconnectivity, in contrast, is linked to depressive and anxiety symptoms,^66^ and is often associated with the emotional distress experienced by individuals with long-term insomnia symptoms.^50^ In the CN +insomnia group, CEN FC alterations revealed an increase in association of all networks with the right LPFC, a key area for the inhibitory control of emotional reactivity.^67^ Persistent hyperconnectivity between the right LPFC and the right parietal DMN showed that this inhibitory control mechanism was still partially present in AD +insomnia; despite a notable hypoconnectivity between the right LPFC and the medial SN, parietal CEN, ventral PFC, and precuneus/PCC. Interestingly, hypoconnectivity between the right LPFC and the left medial SN was also correlated with an increased MMSE score, suggesting perhaps that reduced inhibitory control may be beneficial to daytime cognitive performance. A more notable symptom of AD +insomnia, however, was the significant dysconnectivity of the right parietal CEN with numerous triple network hubs. This dysconnectivity was also reflected in the aggregate DC of the CEN in AD individuals. Decreased FC of the right parietal gyrus is thought to underlie symptomatic anxiety in late-life depression.^66^ Thus, similarly to our findings in the DMN, this dysconnectivity may reflect an exacerbation of comorbid anxiety in AD individuals in the presence of insomnia symptoms.

### Structural Alterations

The present study found no significant alterations in the SC or GMV of any triple network nodes. This finding is in line with current knowledge on (the absence of) morphological changes associated with insomnia symptoms.^51^ However, it also suggests that insomnia symptoms do not significantly accelerate the structural decline present in AD. A possible reason for this could be that the present study investigated acute insomnia symptoms rather than chronic insomnia disorder,^20^ and that significant structural changes might require persistent symptoms.

### Affective Symptoms in AD

Insomnia can come paired with a variety of symptoms that may reflect the mental inability to sleep (i.e. hyperarousal) or the cognitive and affective consequences of (perceived) poor quality sleep (i.e. post-insomnia anxiety). The present study identified correlates of predominantly the latter type in AD individuals with insomnia symptoms. Crucially, cognitive and affective symptoms were strengthened when comparing between AD individuals with and without insomnia symptoms. This may reflect a heightened sensitivity to anxiety or depressive episodes, which is often seen in neurodegenerative disorders. More importantly, however, it reflects a set of phenotypes that may be typically attributed to AD itself. Unfortunately, AD individuals are still rarely screened for insomnia, making it impossible to assess what portion of comorbidities could be associated with insomnia symptoms. Providing treatments such as cognitive behavioral therapy for AD individuals with insomnia could potentially reduce the severity of these comorbidities and improve quality of life.^68^

### Limitations

The present study had some limitations. First, insomnia symptoms were broadly classified because of the limited availability of sleep assessments in the ADNI database. Indeed, the NPI is a more general cognitive well-being questionnaire that is not specifically tailored toward assessing insomnia or sleep disturbances. It also does not assess the frequency of symptoms or daily dysfunction, which are diagnostic criteria for insomnia disorder. In contrast, e.g. the Insomnia Severity Index (Morin et al., 2011) would be able to more accurately capture insomnia symptoms. Utilization of the ISI could also allow further studies to compare the effect of acute and chronic insomnia, the latter of which we did not include in this study. Second, the present study investigated the interaction between insomnia symptoms and AD on cross-sectional data of individuals at distinct stages on the AD continuum. To better assess the long-term interrelationship between the two disorders, large-scale longitudinal datasets are required. Third, the present study was unable to correct the effects of sleep deprivation, which may come paired with insomnia symptoms. With the presence of sleep length as a covariate, either by way of a questionnaire or measurement, future studies could adjust for the effect of sleep deprivation. Finally, the present study suffered from low statistical power at the node level in the main sample, and at all levels in the CSF biomarker subsample. This was primarily due to the large number of exclusions during selection and preprocessing, significantly reducing the *a priori* sample size; the relatively small intersection between rs-fMRI, NPI, and CSF biomarker data on ADNI; and the general low power of multinodal rs-fMRI FC analyses.^69^

### Conclusion

The present study found evidence that insomnia symptoms significantly moderate the effect of AD pathophysiology on the triple network functional organization. Namely, DMN and CEN functional connectivity in individuals with clinical AD and insomnia symptoms is significantly different from cognitively normal individuals with insomnia symptoms. This study posits that insomnia may present atypically in AD individuals and highlights the need for increased screening and therapy for insomnia in AD.

## DATA AVAILABILITY

The Alzheimer’s Disease Neuroimaging Initiative (ADNI) is a multisite initiative for the prevention and treatment of AD. The MRI, CSF biomarker, and clinical data used in the present study is available under the ADNI data sharing policy. Modifications to the data were tracked by DataLad and are reported in a GitLab repository (https://jugit.fz-juelich.de/j.elberse/adni-insomnia).

## Data Availability

The Alzheimer Disease Neuroimaging Initiative (ADNI) is a multisite initiative for the prevention and treatment of AD. The MRI, CSF biomarker, and clinical data used in the present study are available under the ADNI data sharing policy. Modifications to the data were tracked by DataLad and are reported in a GitLab repository (https://jugit.fz-juelich.de/j.elberse/adni-insomnia).

https://adni.loni.usc.edu/

## ACKNOWLEDGEMENTS

The authors thank Debottam Kundu, Max Hinrichs, and Federico Persi for their help with proofreading and statistical analysis.

