## Supplemental file for "The interplay between insomnia and Alzheimer’s Disease across three main brain networks"

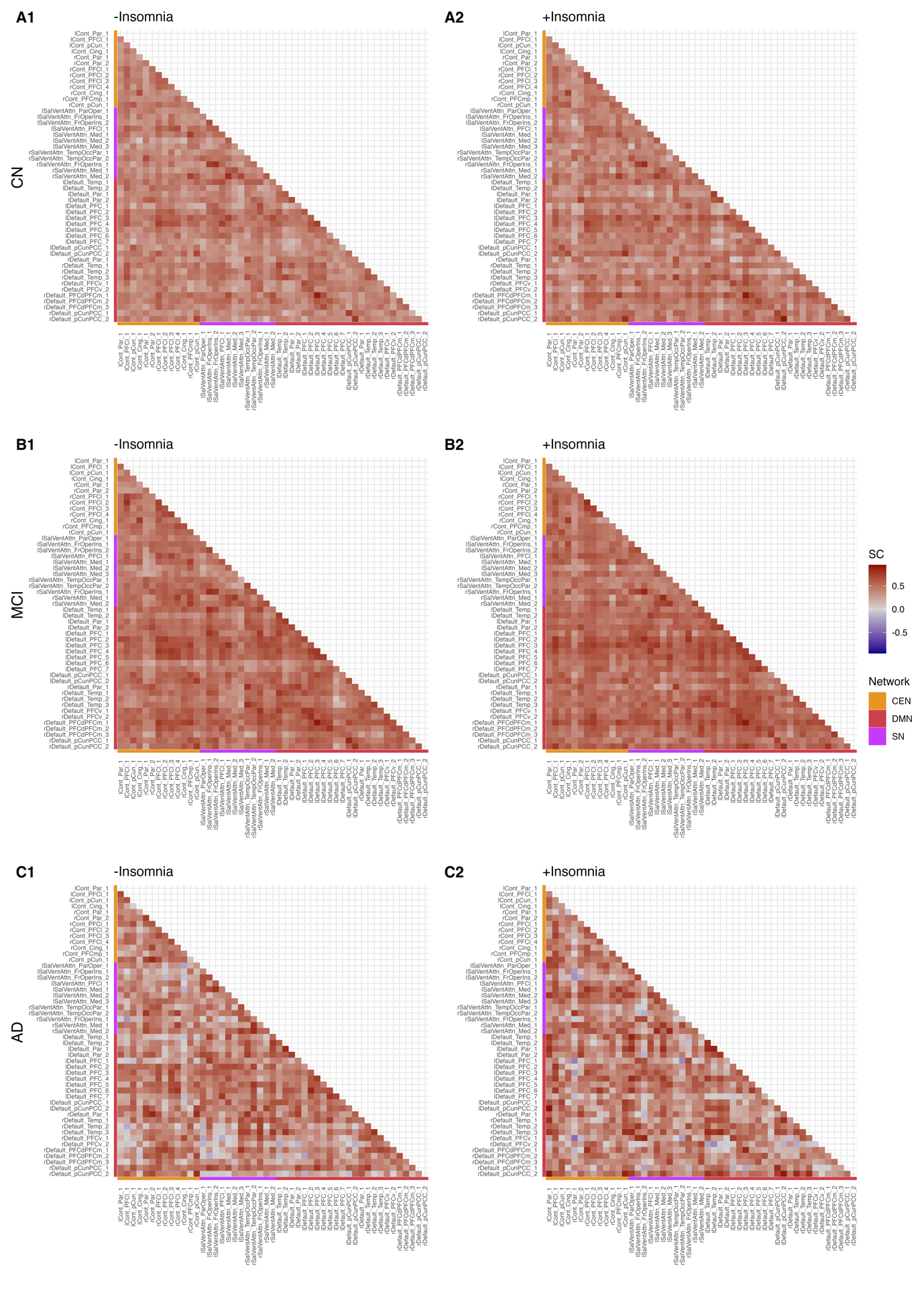

**Figure S1 – Three Network Structural Covariance in the Presence of Insomnia Symptoms.** Node-level alterations in three network SC associated within(**A**) CN, (**B**) MCI, and (**C**) AD, and across the (**1**) -Insomnia and (**2**) +Insomnia conditions. A high SC was observed between nearly all nodes of the triple network. Permutation tests revealed no significant differences between -Insomnia and +Insomnia columns. *CEN = Central Executive Network; Cing = cingulate; DMN = Default Mode Network; Fr = frontal; Ins = insula; Med = medial; Occ = occipital; Oper = operculum; Par = parietal; PCC = posterior cingulate cortex; pCun = precuneus; PFC = prefrontal cortex; PFCd = dorsal prefrontal cortex; LPFC = lateral prefrontal cortex; PFCm = medial prefrontal cortex; PFCmp = medial posterior prefrontal cortex; PFCv = ventral prefrontal cortex; SC = SC; SN = Salience Network; Temp = temporal.*

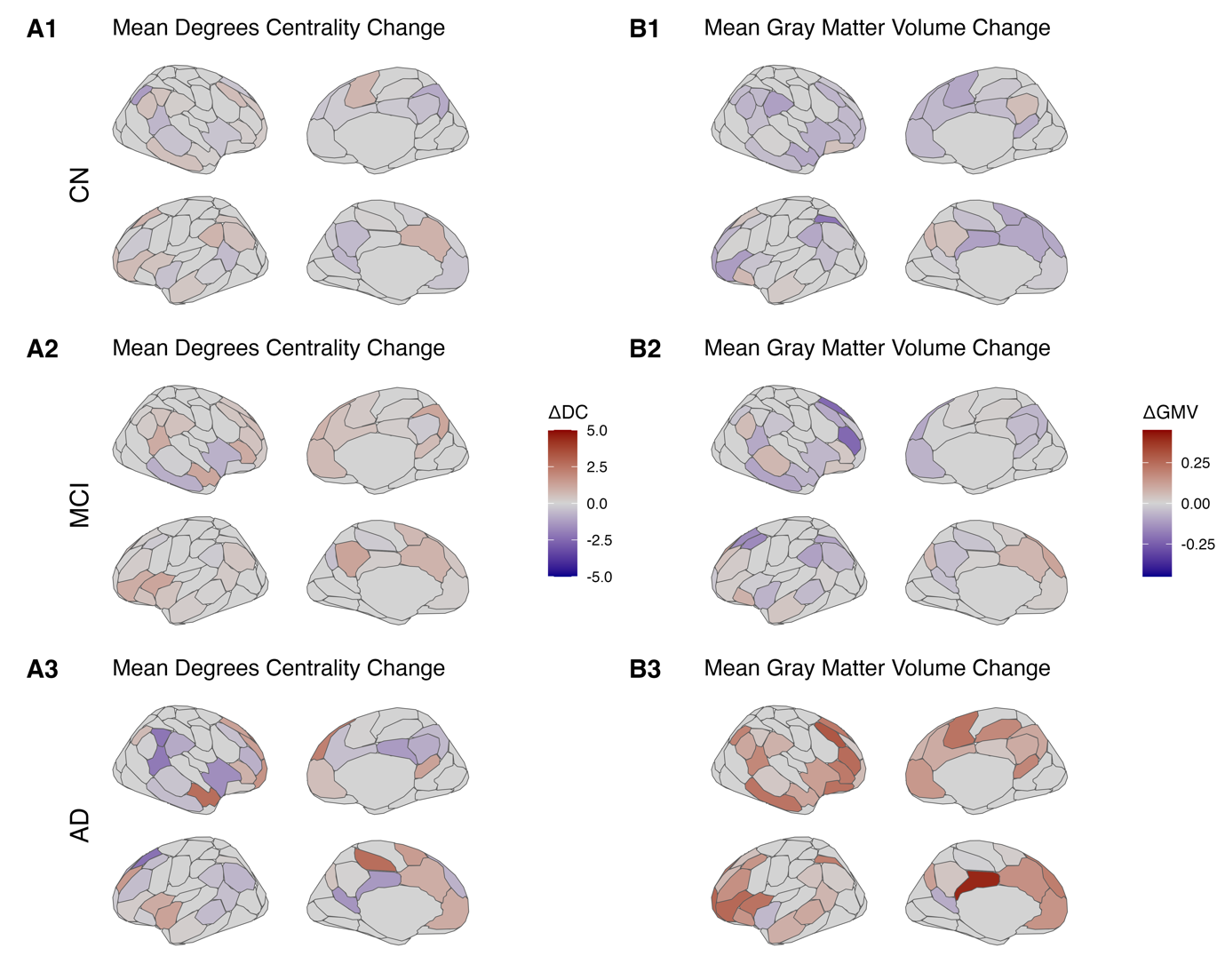

**Figure S2 – Mean Degrees Centrality and Gray Matter Volume Changes associated with Insomnia Symptoms.** Mean node-level (**A**) DC and (**B**) GMV changes associated with insomnia symptoms across (**1**) CN, (**2**) MCI, and (**3**) AD. Insomnia symptoms were associated with various patterns of non-significant DC and GMV changes within each diagnostic group. *∆DC = mean degrees centrality change; ∆GMV = mean gray matter volume change.*

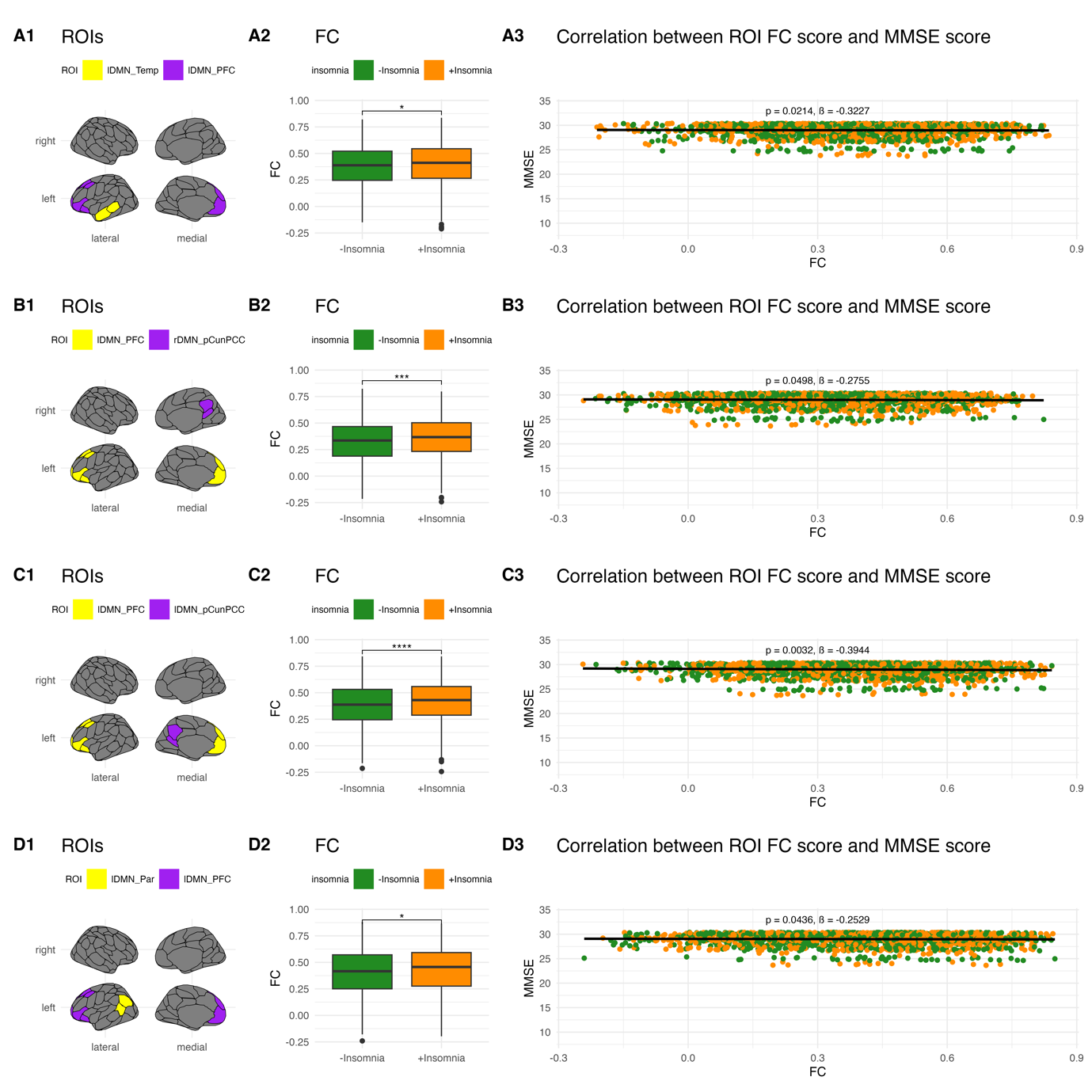

**Figure S3 - Abnormal Inter-ROI Connectivity and MMSE Score in CN.** ROI-level edges in CN +insomnia that predict significant changes in MMSE score. (**1**) ROI-level edges affected by the +insomnia condition. (**2**) Change in FCfor selected edges across the +insomnia condition. (**3**) Overall correlation of ROI-level edge FC scores and MMSE scores. Hyperconnective intra-DMN edges are overwhelmingly associated with decreased cognitive functioning in CN +insomnia. *CEN = Central Executive Network; DMN = Default Mode Network; FC = functional connectivity; Fr = frontal; Ins = insula; Med = medial; Occ = occipital; Oper = operculum; Par = parietal; PFC = prefrontal cortex; LPFC = lateral prefrontal cortex; SN = Salience Network; Temp = temporal. *p < 0.05, **p < 0.01, ***p < 0.001, ****p < 0.0001. ß = estimate.*
